# Characterizing cellular and molecular variabilities of peripheral immune cells in healthy inactivated SARS-CoV-2 vaccine recipients by single-cell RNA sequencing

**DOI:** 10.1101/2021.05.06.21256781

**Authors:** Renyang Tong, Jianmei Zhong, Ronghong Li, Yifan Chen, Liuhua Hu, Zheng Li, Jianfeng Shi, Guanqiao Lin, Yuyan Lyu, Li Hu, Xiao Guo, Qi Liu, Tian Shuang, Chenjie Zhang, Ancai Yuan, Minchao Zhang, Wei Lin, Jun Pu

**Author notes:** To whom correspondence should be addressed: Jun Pu, MD, Ph.D.; State Key Laboratory for Oncogenes and Related Genes, Division of Cardiology, School of Medicine, Renji Hospital, Shanghai Cancer Institute, Shanghai Jiao Tong University, Shanghai, China., Wei Lin, Ph.D.; State Key Laboratory for Oncogenes and Related Genes, Division of Cardiology, School of Medicine, Renji Hospital, Shanghai Cancer Institute, Shanghai Jiao Tong University, Shanghai, China. These authors contributed equally to this work (Renyang Tong, Jianmei Zhong).

## Abstract

We systematically investigated the transcriptomes of the peripheral immune cells from 6 inactivated vaccine, BBIBP-CorV recipients at 4 pivotal time points using single-cell RNA-seq technique. First, the significant variation of the canonical immune-responsive signals of both humoral and cellular immunity, as well as other possible symptom-driver signals were evaluated in the specific cell types. Second, we described and compared the common and distinct variation trends across COVID-19 vaccination, disease progression, and flu vaccination to achieve in-depth understandings of the manifestation of immune response in peripheral blood under different stimuli. Third, the expanded T cell and B cell clones were correlated to the specific phenotypes which allowed us to characterize the antigen-specific ones much easier in the future. At last, other than the coagulopathy, the immunogenicity of megakaryocytes in vaccination were highlighted in this study. In brief, our study provided a rich data resource and the related methodology to explore the details of the classical immunity scenarios.

## Introduction

As of May 2021, World Health Organization (WHO) has reported 148 million Coronavirus disease 2019 (COVID-19) infections and 3.1 million deaths worldwide (https://covid19.who.int/). Promotion and popularization of vaccine is vital to disease control, with its safety and efficacy of the top priority. There are multiple SARS-CoV-2 vaccines available and several vaccines under development, including the inactivated, genetically engineered subunit, adenovirus and nucleic acid vaccine types. So far, a number of institutions of different countries and regions have reported the results of safety and immunogenicity of SARS-CoV-2 vaccine on phase III clinical trials or phase III interim analysis, such as, vaccines BNT162b2^1^ and mRNA-1273^2^, adenovirus vaccines AZD1222^3^, Ad5-nCoV^4^ and inactivated vaccines BBIBP-CorV^5^ and CoronaVac^6, 7^, and protein subunit vaccine NVX-2373^8^. For example, Ugur Sahin *et al*.^9^ demonstrated that RNA vaccine (BNT162b2) not only elicited adaptive humoral, but also stimulated the cellular immune response even at the low-dose vaccine. The new vaccine types such as adenovirus-vectored vaccines and RNA vaccines have emerged and fast developed with good efficacy in recent years, though severalserious adverse events have been reported^10, 11^ ^12, 13^. Their efficacy and long-term safety were yet to be reviewed by a follow-up study. With the support of long clinical practice, traditional inactivated vaccines play an irreplaceable role in the prevention and control of infectious disease. As a pattern of inactivated SARS-CoV-2 vaccines, BBIBP-CorV, was manufactured by Sinopharm China National Biotec Group (CNBG) and contained a SARS-CoV-2 strain inactivated inside Vero Cells^14^, which was confirmed for its safety, tolerability, and immunogenicity in clinical trials of healthy people in China^5^ and UAE, Bahrain, Egypt and Jordan (https://cdn.who.int/).

Single-cell RNA-seq (scRNA-seq) technique has demonstrated its power in the research of cell biology by deconvoluting the transcriptional signals in the multi-cellular samples of heterogenous cellular compositions^15,16^. Other than characterizing the cell types in a heterogeneous tissue, it enables us to detect the variabilities within a cell lineage. The recent pandemic of COVID-19 has prompted the demand of single-cell studies in order to uncover the characteristics immune responses of different immune cell types during the symptom development of COVID-19^17, 18, 19, 20^. These published scRNA-seq datasets provide abundant and detailed reference information of this disease, though, no results have been reported for any COVID-19 vaccines yet. Moreover, a comprehensive comparison of the human immune responsive signals across the transcriptome at single-cell resolution over the time becomes practical and necessary. With no doubt, such detailed comparison facilitates the in-depth understanding of the pathogenesis, symptom development and immunization of COVID-19 which guides us to the better vaccination strategy. Therefore, we aimed to look through a ‘sight glass’ of the human immunity system, a.k.a., peripheral blood mononuclear cells (PBMCs) to monitor the manifestation of vaccine response of the canonical immune cells. In the meantime, we could take the advantage of the recent published scRNA-seq datasets of COVID-19 patient samples^21^ and Flu vaccination samples^22^ to achieve such a goal. Besides, the roles of megakaryocytes/platelets in COVID-19 recently have attracted some attentions due to recent reports of blood-clot side effect by AZD1222^12, 23^. Megakaryocytes were also found to contribute to the complication of COVID-19 patients and be correlated to the organ failure and mortality^24, 25, 26^. We were therefore also interested in looking into the signals related to immune-responsive and coagulopathy in this non-canonical immune cell type using the scRNA-seq dataset.

In this study, we extensively investigated the PBMC samples of BBIBP-CorV vaccine recipients. Our scRNA-seq data delineated a high-resolution transcriptomic landscape of peripheral immune cells during the vaccination of a classical vaccine type. Meanwhile, we integrated the PBMC scRNA-seq datasets of COVID-19 patients and flu vaccination in order to identify the common and distinct variation trends among our data set and the two other data sets. All these efforts could provide a rich data resource and demonstrate a methodology to explore the human immunity in disease development and vaccination at great details. Moreover, this work could facilitate the in-depth understandings of the manifestation of immune response in peripheral blood under different stimuli and provide many clues to design the new assay for the assessment of the immune protection and adverse effects after the vaccination or infection recovery.

## Results

### Overview of immune responses and peripheral immune cells variations by scRNA-seq

To assess the immune cells content and the phenotypic alterations following the vaccination with inactivated SARS-CoV-2 vaccine (BBIBP-CorV), six healthy young adults with 4 males and 2 females from Pu Dong Cohort, China, were enrolled in this 2019-nCoV vaccine observational study [NCT04871932] according to inclusion and exclusion criteria (Fig.1A). All six volunteers received 2 doses of inactivated SARS-CoV-2 vaccine (BBIBP-CorV). Blood samples were collected at 4 time points including the day before vaccination (BV), 7 days after first dose (1V7), 7 days after second dose (2V7) and 14 days after second dose (2V14) (Fig. 1A). Then, serum and PBMCs were separated from all time points of samples within 2-hour of collection. The 2019-nCoV Spike RBD antibodies and cytokines were measured in serum. After isolating the PBMCs, we profiled V(D)J repertoires of T and B cells integrated with 5’ Gene Expression using droplet based single-cell sequencing method (10× Genomics) (Fig.1A).

**Figure 1.**
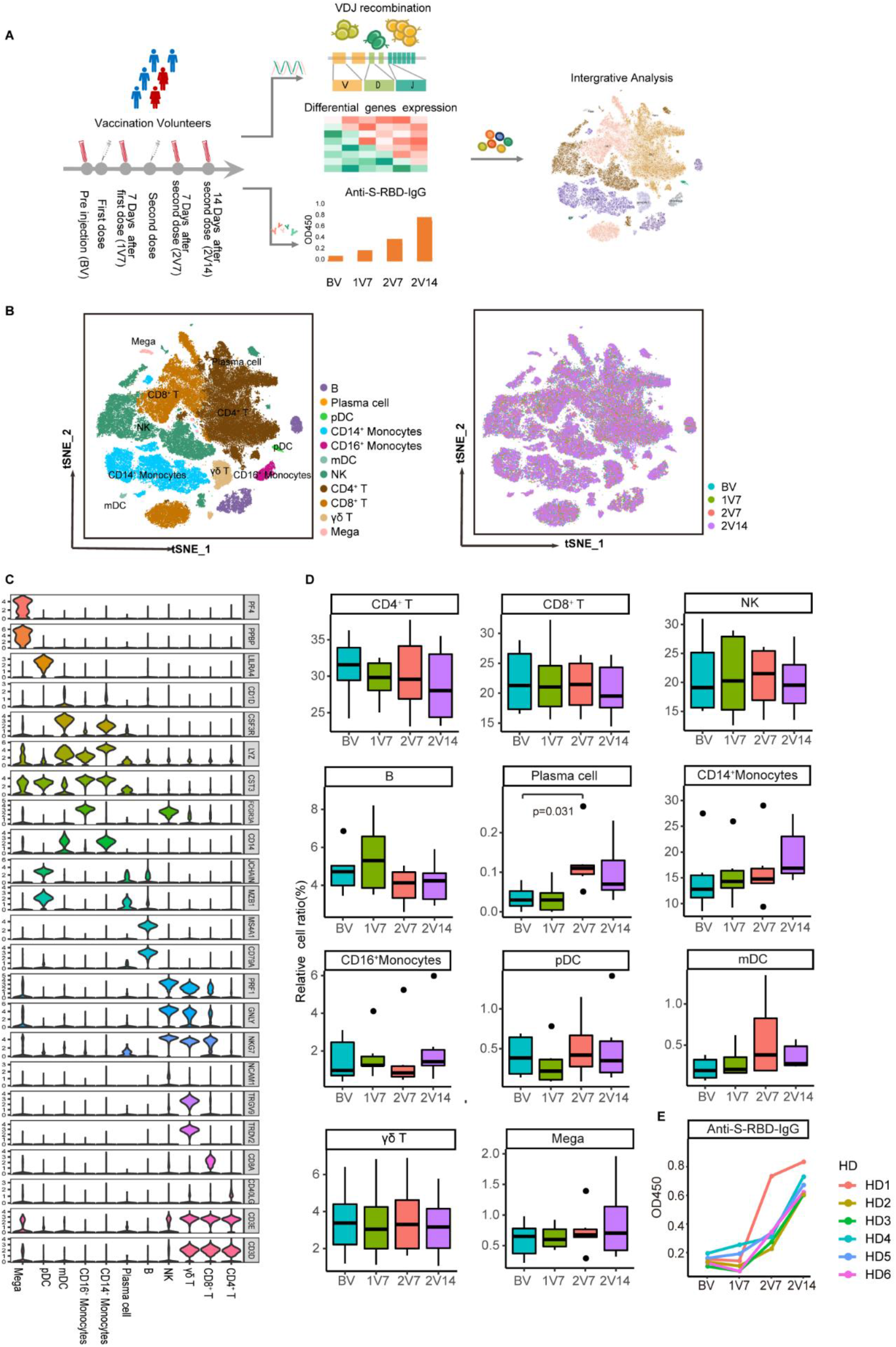
Overview of vaccination efficacy, scRNA-seq data quality, cell compositions. **A**. Workflow of vaccination, sampling and scRNA-seq; **B**. tSNE charts of ∼120 k single cells by color coded by putative cell types and time points; **C**. Markers for the putative cell types using violin plots in scRNA-seq data; **D**. Box charts showing variabilities of the contents of each cell type by time point. All differences with p < 0.05 and analyzed by two-sided Student’s t-test; **E**. Serological responses of 6 healthy recipients to recombinant S-RBD at 4 time-points. Dilution of 1:55 for IgG.

Based on the integrated single-cell gene expression profiles, we recovered the transcriptomes of ∼120k high-quality cells in total from 24 samples and the total cells were combined and projected in a 2-D space using the t-distributed stochastic neighbor embedding (t-SNE). Unsupervised clustering analyses and marker mapping enabled us to uncover 11 major types (Fig.1B). The markers for the 11 major immune cell lineages were shown on Fig.1C. As the shown in Fig.1D, summary statistics of the 11 cell-type fractions showed plasma cells increase significantly after the second dose, whereas the other 10 cell types pretty much remained the same level.

Previously, Wang *et al*.^14^, Xia *et al*.^5^ have already demonstrated the satisfactory efficacy by increasing seroprotective antibody titers in the animal experiments and human clinical trials by this vaccine type. To further validate the efficacy in our participants, we measured 2019-nCoV Spike RBD antibodies using indirect enzyme-linked immunosorbent assay (ELISA) kits. Anti-S-RBD-specific IgG antibody level significantly increased after the second dose for total 6 participants (Fig.1E). The growth of plasma cells content and the observable production neutralizing antibody by second dose suggests the robust humoral immune response. The cellular immunity was also assessed. 10-cytokine serum assays were first performed on the total 24 samples on bead-based flow-cytometric assays on the BD LSRFortessa X-20 platform, though, no obvious post-vaccination increase was observed (Fig.S1A). We then used an ultra-sensitive bead-array-based immunoassay (single-molecule array technique) to validate the IL-4 signal and found that the quantification was consistently lower (one order of magnitude) than the readings of BD LSRFortessa X-20 platform (Fig.S1B), which suggested, 1. most of the cytokines remained low level at these time points. 2. limit of quantification of bead-based flow-cytometric assays likely above the real concentrations of these cytokines in these samples (Fig. S1B).

### Classical immune responsive signals of major immune cell types in vaccination

After the successful characterization of the major cell types, we were interested in investigating the gene expression patterns in the major immune cell types. The differentially expressed genes (DEGs) were first identified in 11 cell types by the comparisons between the post-vaccination samples vs. the pre-vaccination samples (BV). The DEGs list was shown in Supplementary table 1. The unsupervised clustering of these DEGs exhibited a pattern of 3 dominant clusters for the 24 samples in most of the cell types. We used the heatmap of B cells as an example to demonstrate the sample clusters (Fig.2A). The red cluster included the samples mostly in the 2V7 or 2V14 time points, which reflected an active immune responsive state (R-state). The green cluster included the samples mostly in early BV or 1V7 time points, which likely reflected the baseline or minor-responsive state (B-state). The blue cluster covered broader time points, which likely reflected a post-vaccination protection state (P-state) or recovery state. The identical clustering patterns were observed when the regulons heatmap were calculated and plotted using SCENIC^27^ (Fig.2B). Such unsupervised clustering provided an unbiased sample grouping. More details of the signal variabilities by heatmaps for the major 6 cell types were shown in Fig.S2A-2J.

**Figure 2.**
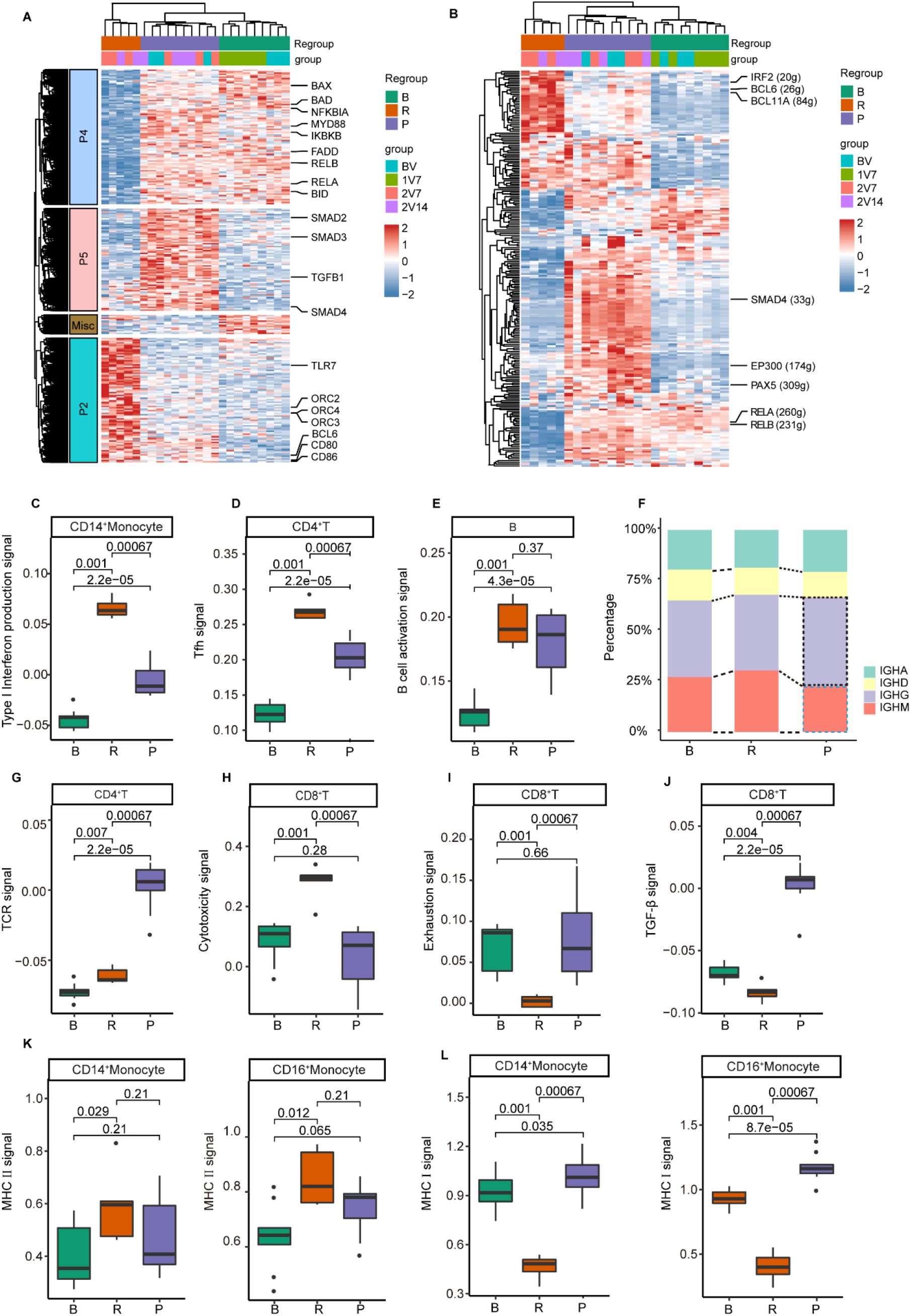
Classical immune responsive signals of major immune cell types in vaccination. **A**. Heatmap of DEGs detected in B cells; **B**. Heatmap of regulons in B cells; **C**. Modular scores of type I interferon production related signals; **D**. Tfh signal of CD4^+^ T cells; **E**. B cell activation signal of B cells; **F**. Relative UMI percentages of different classes of antibody genes; **G**. TCR signal of CD4^+^ T cells; **H**. Cytotoxicity signal of CD8^+^ T cells; **I**. Exhaustion signal of CD8^+^ T cells; **J**. TGF-β signal of CD8^+^ T cells; **K**. Type II MHC signal of Monocytes; **L**. Type I MHC signal of Monocytes. All differences with p < 0.05 and analyzed by two-sided wilcox test.

The DEGs were re-calculated based on the updated sample clusters (B-R-P) and the following 15 variable signaling keywords were identified based on these DEGs. The updated DEGs list for this comparison was shown in Supplementary table 2 and showed very little change. The coordinated gene expressions of a certain functional group or signaling pathway suggested their involvement in the response to BBIBP-Corv vaccination. We used simple schematic diagrams to illustrate the 5 dominant patterns of signal variation observed in the heatmaps to delineate the variability of immune-responsive signals in B-R-P state as Fig.S2K.

As expected, we observed the elevation of the monocytes’ type I interferon production signal (Fig.2C, pattern 2), which was an essential signal of innate immunity for viral suppression. We also observed significant elevation of the antigen-specific follicular T helper cell (Tfh) activation (*CD40LG, ICOS, SLAMF1*, etc.) (Fig.2D, pattern 2) and the B cell activation signal (*TLR7, CD80/CD86, BCL6*) (Fig.2E, pattern 2), that were associated with neutralizing antibody production. In the meantime, we evaluated the expression of different classes of constant gene of Immunoglobulin heavy chain. The IGHG went up and IGHM went down in protection state (P-state), suggesting the occurrence of class switching of neutralizing antibody molecules (Fig.2F).

TCR responsive signals (*VAV3, AKT3, MALT1, MAPK1*, etc.) (Fig.2G, pattern 1), as well as cytotoxic signal were also enhanced (Fig.2H, pattern 3). These lymphocyte-related signals contributed to antigen-specific immunity, a.k.a., adaptive immunity.

The immune regulatory signals such as TGF-β pathway (*TGFB1, TGFBR1/2, SMAD2/3/4*, etc.) (Fig.2J pattern 5), and T cell exhaustion (*LAG3, PDCD1*) (Fig.2I pattern 4) were suppressed in the high immune-responsive samples (R-state) until they went up to play their roles in the protection state (P-state).

We also identified some changes contradictory to the expectation. For example, NF-κB signal was expected to be upregulated in the most of the immunity situation (Fig.S2L). However, NF-κB signal actually went down (pattern 4). Second, instead of enhancing the antigen presenting, MHC I signal significantly dropped in most of the peripheral immune cell types in R group (Fig.2L, pattern 4). Though, type II MHC genes mostly went up as expected (Fig.2K, pattern 2). Later, when we examined COVID-19 dataset^21^, we didn’t observe such changes in COVID-19 patients. Why this change occurs and what outcomes will this change brings are elusive.

Other variabilities of the classical immune-responsive signals including cell-cycles and apoptosis (Fig.S2M-2N) were summarized in Supplementary table 3. High cell-cycle signal (*ORC2, CCNH, CDC7, CDK7*, etc.) and low apoptosis signal (*BAX, BAD, TP53*, etc.) indicated the high proliferative potential of the cells in R-state samples.

### Characteristic transcriptions of the top TCR and BCR clones

As our scRNA-seq study also included TCR and BCR V(D)J sequencing results, it allowed us to track down the TCR/ BCR clonal expansion in these samples. The results showed the diversities of the T cell and B cell clones wend down after vaccination due to the expansion of the antigen-specific clones (Fig.3A, 3D and 3G). The top 20 abundant clones of both T cells and B cells were pulled out to investigate their characteristics in comparison with the low abundance clones as the counterpart.

**Figure 3.**
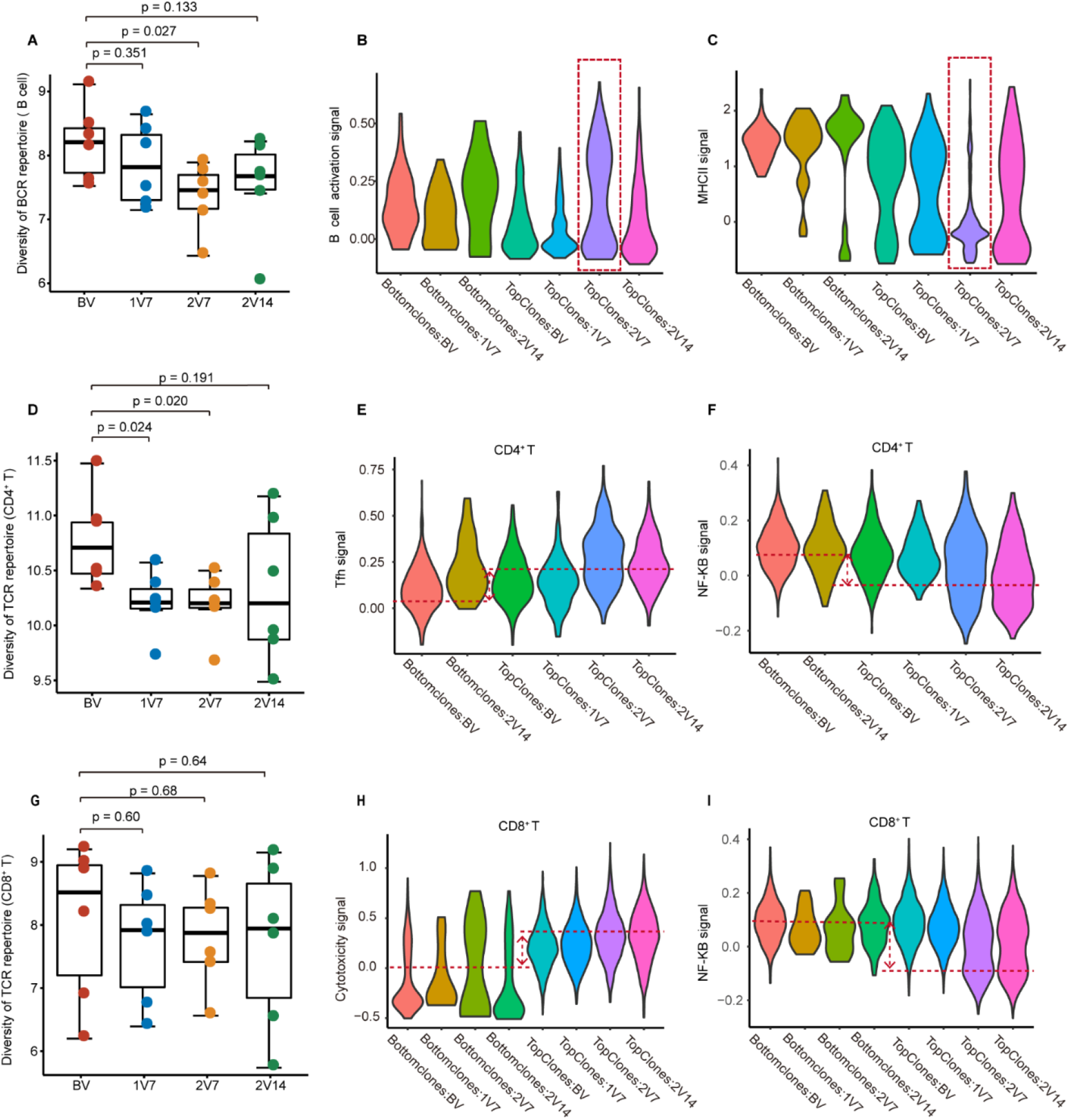
Characteristic transcriptions of top TCR and BCR clones. **A**. Diversity of BCR repertoire at different vaccination time points; **B** and **C**. variable immune-responsive signals in top BCR clones; **D** and **G**. Diversity of TCR repertoires of CD4^+^ T and CD8^+^T cells at different vaccination time points; **E** and **F**. Variable immune-responsive signals in top CD4^+^ T cells TCR clones; **H** and **I**. Variable immune-responsive signals in top CD8^+^ T cells TCR clones. All differences with p < 0.05 and analyzed by two-sided Student’s t-test.

First of all, it was found that top BCR clones were enriched in the samples after second dose, indicating the observable clone expansion occurred after the second dose. Second, it was found that the top 20 clones were highly correlated to low MHC II signal and high B cell activation signal (Fig.3B and 3C). This was expected, though, it was still encouraging to confirm that the possible antigen-specific B cells could display transcriptional signature in the peripheral blood, which made it more specific to access the antigen-specific BCR sequence using single-cell sequencing technique.

Top TCR clones didn’t necessarily occur in the post-vaccination time, which made it less straightforward to confirm their vaccine-specificity. We were still able to find the correlation between the top T cell clones and the immune-responsive signals. In CD4^+^ T cells, it was found that the top clones were associated with high follicular T helper (Tfh) and low NF-κB signal (Fig. 3E and 3F). In CD8^+^ T cells, it was found that the top clones were associated with high cytotoxic but low NF-κB signal (Fig.3H and 3I).

### Non-trivial roles of megakaryocytes in vaccine immunity

Megakaryocytes were mostly recognized for its role in the hemostasis and wound healing. Little has been discussed for its contribution in the disease or vaccine immunity. After examining this atypical immune cell type, we observed the gene expression variation of megakaryocytes in vaccine immunity.

First, the variation of platelet aggregation signal (*GP9, ITGB3, TBXA2R, FLNA, VASP*, etc.) in megakaryocytes (Fig.4A and 4B, pattern 4) were identified. These genes were significantly down-regulated in the immune-responsive samples (R-state), which suggested low risk of thrombosis during the period of BBIBP-CorV vaccination. Later, when we inspected the same set of genes in the COVID-19 patients^21^ and influenza vaccination^22^ samples at the corresponding immune-responsive time point, instead of getting down-regulated, we found the platelet aggregation signal significantly went up (dashed red box in Fig.4E and Fig.S3D). That means, the variation of platelet aggregation signal could be distinct by different vaccines.

As a myeloid-lineage cell type, it was found that megakaryocytes actually contributed more inflammatory signal than we thought, such as interleukin-1 beta (IL-1β) production (*TLR4, IFI16, CASP1, CASP8, NLRP1, NLRP2*, etc.) and neutrophil activation (*SA0018, S100A12, FCER1G, VNN1*, etc.) (Fig.4A, 4C and 4D). The expression levels of the related genes were actually as high as the monocyte population (Fig. S2O and S2P), which was considered the dominant cell type of innate immunity in PBMCs. In flu vaccination, we observed the elevation of IL-1β production and neutrophil activation signal as well at 28-day sample, the putative high response time point (Fig. 4F and 4G). All these results suggested the roles of megakaryocytes in pathogenesis and vaccination should not be overlooked.

**Figure 4.**
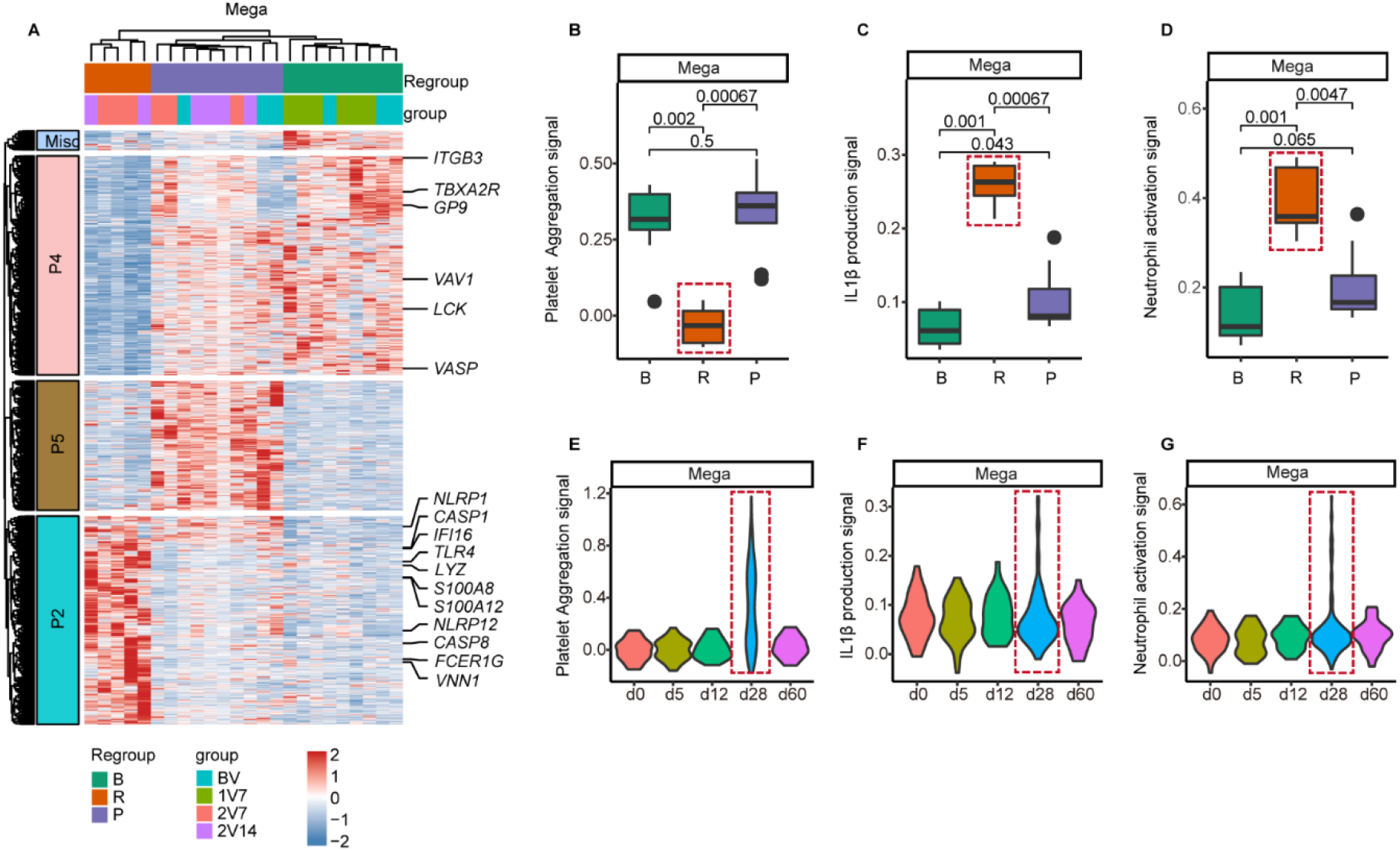
Non-trivial roles of megakaryocytes in vaccine immunity. **A**. Heatmap of DEGs detected in megakaryocyte**s; B**. Platelet aggregation signal of megakaryocytes; **C**. IL-1β production signal of megakaryocytes; **D**. Neutrophil activation signal of megakaryocytes; **E-G**. Platelet aggregation signal, IL-1β production signal and Neutrophil activation signal of megakaryocytes at 28-day of flu vaccination. All differences with p < 0.05 and analyzed by two-sided wilcox test.

### Transcriptomic similarity and distinction of COVID-19 and vaccination immunity in PBMC

It was postulated that the dysregulated immune response could be reflected in differential signals between the COVID-19 infection patients and the vaccine recipients. We were particularly interested in looking for the signatures that were associated to the symptom development and the later immune protection. By examining the similarity and the distinction of the developmental trend of these signals, we aimed to gain more insights of both disease development and vaccine protection.

We used the keywords in Supplementary table 3 to assess the COVID-19 samples. There were three natural sample groups in previous COVID-19 study, i.e., healthy control, progression samples of mild and severe symptoms and the convalescent samples of both patient types^21^. Other than the resemblance between COVID-healthy samples and the vaccine-baseline samples, it was intuitively believed that the COVID-convalescence samples were similar to the vaccine-protection samples (P-state). What do these immune signals vary in the progression samples are of our interest.

Many of the abovementioned signals only showed an obvious turning point at convalescence group, including type I interferon productions, Tfh and B cell activation, TGF-β, cell-cycle, apoptosis and so forth (Fig.5A. and Fig.S3A-S3B). We were able to observe obvious signal boundary line between healthy/progression samples and convalescence samples. That means, unlike what happened in vaccination, some of the critical signals of immunity were not initiated but suppressed until the recovery.

**Figure 5.**
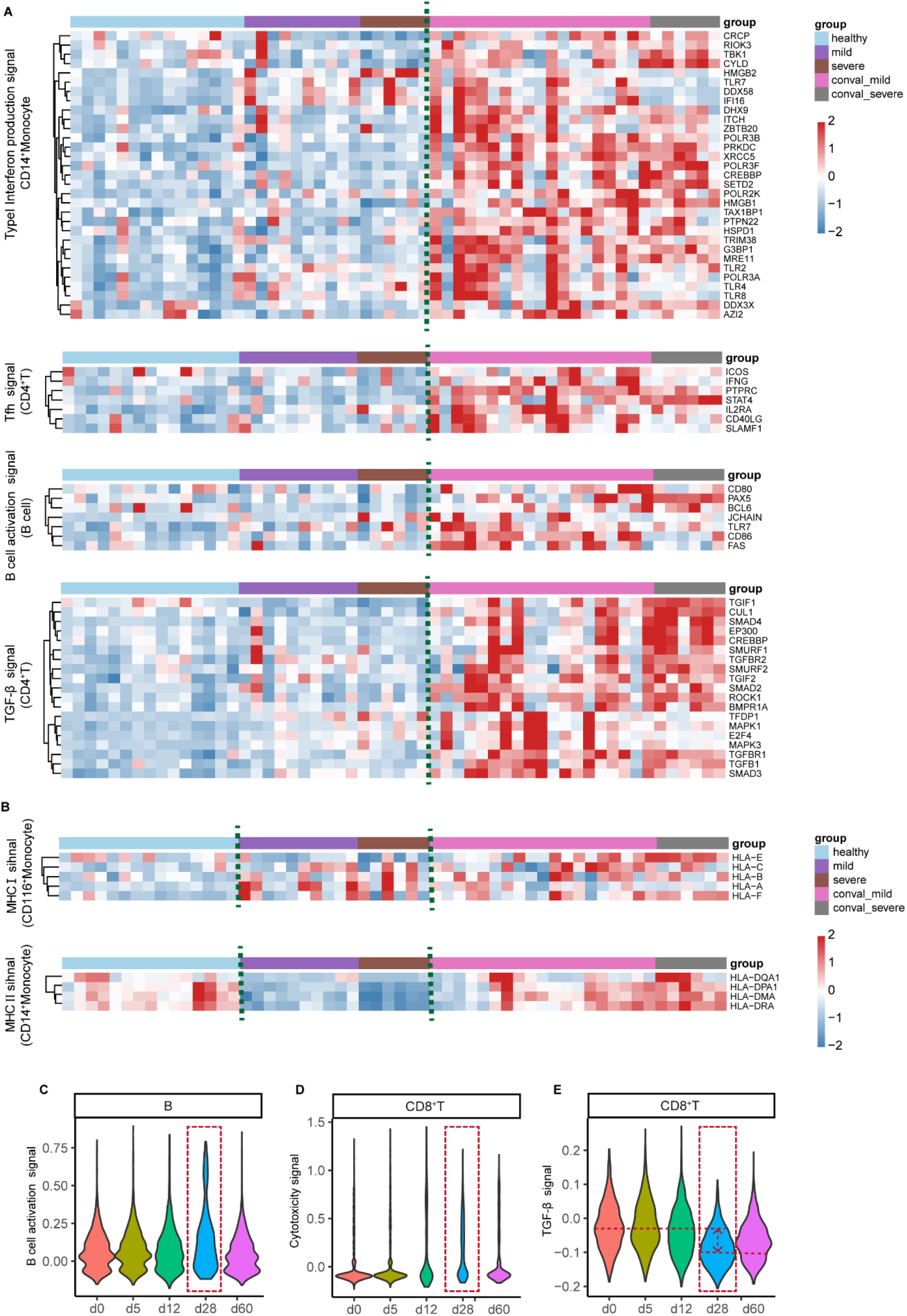
The immune-responsive signals in COVID-19 patients and Flu vaccination. **A**. Turning point of immune-responsive signals at convalescence; **B**. Variability of type I and type II MHC signals at COVID-19 patients. **C-E**. Variable immune-responsive signals at 28-day of flu vaccination.

In general, there were many immune responsive signals in certain cell types in the progression samples whose trends were similar to those in R-state of vaccination. For examples, cytotoxic signal in T cells and NK cells significantly increased in the patients’ progression samples consistent with Zhang *et al*. and Wilk *et al*. study^17, 28^ (Fig.S3C).

We also revealed the distinctions between disease immunity and vaccination immunity using both datasets, especially for type I and type II MHC. In those antigen presenting cells like B cells and monocytes, type I MHC signal significantly decreased in vaccine response group (R group) whereas it tended to increase in the progression patients (Fig.5B). Type II MHC signal was suppressed in progression in both mild or severe patients (Fig.5B), whereas it was up-regulated during the vaccination. Another distinction was that T cells exhaustion signal went down in vaccination but went up in progression patients (Fig.S3C).

In summary, we identified the distinct signal variation patterns between the COVID-19 progression and vaccination samples, such as 1. type I and II MHC signals 2. T cells exhaustion signal.

### Potential effects of the identified variable classical immune signals in flu vaccination

To further evaluate the responsive role of the variable classical immune signals we had identified in inactivated SARS-CoV-2 vaccination, a specific comparison was adopted with a public scRNA-seq data set on the lymph nodes and PBMC samples of the quadrivalent influenza vaccination^22^. This data set was designed for a single-dose vaccine and included 5 time points of PBMC samples with a long post-vaccination follow-up time (2 months).

Using the 15 variable signaling keywords identified in BBIBP-Corv vaccination and the involved differential genes, we calculated their modular scores for each cell in flu dataset. The signal curves showed that 28-day sample gave the most obvious variation in response to the flu vaccine compared to other time points. We were able to detect high B cell activation signal in B cells (Fig.5C), high cytotoxic signal in CD8^+^T cells (Fig.5D), and low TGF-β signal (Fig.5E) in most of the immune cells at 28-day time point.

Elevated B cell activation, elevated cytotoxic signals and suppressed TGF-β signal at 28 days suggested this sample was mostly corresponding to the COVID-19-vaccine samples at immune R-state. We were not able to detect the variation of other signature signals, possibly because their sampling missed the pivotal time point on this volunteer. The megakaryocytes also had shown the significant variation at the same time point, which was mentioned earlier (Fig.4E).

## Discussion

In this study, we extensively investigated the variability of the single-cell transcriptomes of the peripheral immune cells in the process of vaccination by a classical inactivated viral vaccine. Single-cell sequencing technology enabled us to deconvolute the immune-responsive signals into specific cell types.

Unsupervised clustering on the cell types from these samples defined three dominant clusters of the immune cells that were putatively corresponding to different immune-responsive states. Such an unbiased method overcame the challenge of sample grouping by the distinct immune-background, vaccine susceptibility or variable pace of immune responses by different participants.

The coordinated up-/down- regulation of the gene expressions of a certain signaling pathway suggested their involvement of the immune response during the vaccination. Tracking down the variation of these signals in a typical vaccination process allowed us to uncover their possible roles in the immune protection. Using this method, we observed the activation of both humoral immunity and cellular immunity by BBIBP-CorV, including cytotoxicity, follicular helper T mediated immunity as well as B cell activation *et al*. Meanwhile, our results provided a good reference for the future study of other vaccine types.

Surprisingly, we observed the down-regulation of NF-κB signal in the post-BBIBP-CorV vaccination PBMC samples. Previous study suggested that the suppression of NF-κB signal likely alleviated the vaccine adverse reaction^29, 30^, which might partially explain much less adverse reaction cases was reported by the vaccine recipients of BBIBP-CorV. Even though, the antibody productions of our participants were satisfactory.

The correlation between the top TCR/BCR clones and the modular scores of the keywords reveals what phenotypes have been associated with the clones of expansion. Some of the signature transcriptions identified in this work enabled us to capture the target B cells for full-length sequencing and molecular cloning of the antigen-specific antibodies.

The platelet aggregation signal and inflammatory signals were also inspected in the megakaryocytes due to the rising concern on the blood-clot side-effects reported on several vaccine types^10, 11, 12^. This cell type was usually overlooked in immunity, though, in response to vaccine, the inflammatory gene expressions in megakaryocytes not only commensurate with the monocytes but also in a coordinated fashion. In addition, unlike severe COVID-19 samples and influenza vaccine samples, our result showed a down-regulation of blood-clot-related gene expression in our samples under active response (R-state), which likely explained the much fewer reports of thrombus-related adverse events by BBIBP-CorV vaccine^5^.These findings necessitate a more extensive investigation on the roles of the megakaryocytes in the human immunity in the future.

The comparison of the immune-responsive signals between the COVID-19-progression and the vaccination allowed us to identify the discrepancy of immune responsive signals and made it possible to locate the missing puzzle pieces of the unsuccessful or incomplete immunity that contributed the symptoms in patients. These signaling pathways could be used to assess the immunity or as the target of the intervention for the control of COVID-19 symptoms or the vaccine adverse reaction. Though, more validation experiments will need to be done in the future.

We also inspected these signals using the dataset generated from a single-dose flu vaccine recipient^22^. The results showed a significant fluctuation, especially, the B cell activation signal displayed a peak at 28-day. This was concordant to the detection of high content of CD20^hi^CD38^int^ B cells from lymph nodes at 28-day in the original paper, though, a single case study might limit the power of this study.

Our study has some limitations. Due to the currently high cost of single-cell sequencing and inaccessibility of other vaccine type at the moment, the coverage and the size of sampling of this study are relatively limited, which limits the power of this study. Due to the urgent need of COVID-19 related research, the follow-up time couldn’t be long enough, either.

Despite of these limitations, we were able to observe the protective immunity induced by BBIBP-CorV at the single-cell level and identified a set of genes that not only featured the immune-responsive variation of the peripheral immune cells but also exhibited many interpretable signaling pathways. The signature expression described in here might facilitate to design a new assay to assess the immune protection and adverse effects after the vaccination or infection recovery.

## Materials and Methods

### Participants and ethics

All human samples used in this study were processed under Institutional Review Board approved protocols at Shanghai Jiao Tong University. All study participants were recruited after providing informed consent and with approval by the Ethics Committee of Ren Ji Hospital (KY2021-046), School of Medicine, Shanghai Jiao Tong University, and the study was conducted according to the criteria set by the Declaration of Helsinki^31^ (2013). This study was registered with ClinicalTrials.gov (NCT04871932). A written informed consent letters were routinely obtained from all participants in the study and all relevant ethical regulations regarding human research participants were complied with by the investigators. Six healthy non-frail individuals who had not received any vaccine in the past one year were recruited in the Pu Dong Cohort, China, including 4 males and 2 females, aged 30-40 years old. All participants were confirmed negative result of SARS-CoV-2 infection by RT-qPCR tests and negative result for serum SARS-CoV-2-specific IgM/IgG-antibody ELISA tests. The participants had no history of epidemiological contact of COVID-19 patients. They were free of clinical symptoms including fever, cough, headache, sore throat, malaise, loss of smell, runny nose, abdominal pain, diarrhea, joint pain, wheezing or dyspnea within 28 days before vaccination; They were also free of any steroid usage, chronic diseases, pregnancy, lactation, and obvious allergy to any known ingredients contained in the inactivated vaccine.

### Vaccine and Vaccination protocol

The inactivated SARS-CoV-2 vaccine (Vero cells), a single-dose schedule of 4μg/0.5ml, were supported by the Beijing Institute of Biological Products (Beijing, China). Vaccine recipients received BBIBP-CorV twice according to drug instruction via intra muscular with 2 to 4 weeks between each injection.

### Sample collection, preparation and storage

Peripheral venous blood samples were collected at 4 time points including the day before vaccination (BV), 7 days after first dose (1V7), 7 days after second dose (2V7) and 14 days after second dose. The blood samples were processed within 2 hours of collection. The peripheral blood mononuclear cells (PBMCs) were isolated using Vacutainer CPT tubes (BD) according to the manufacture protocols^32^, and cells were immediately used 10× single-cell RNA-seq or cryopreserved in 10% dimethylsulfoxide in FBS and stored at −80°C freezer. The viability and quantity of PBMCs in single-cell suspensions were determined using Trypan Blue. The cell viability of total samples is > 90%. Blood samples in the coagulation promoting tubes were set upright on the tube stand for 20 minutes at 4 °C and were centrifuged at 1000g for 10 minutes at 4 °C. The supernatant serum samples were extracted and stored at −80 °C for future assays.

### Antibody and Serum cytokine detection

The levels of anti-SARS-CoV-2 (2019-nCoV) Spike RBD IgG in serum were tested using Sino Biological® SARS-CoV-2 (2019-nCoV) Spike RBD Antibody Titer Assay Kit (Catalog Number: KIT002) following the manufacturer’s instructions. Absorbance was read at 450 nm with a subtraction at 570 nm by MultiSkan FC reader (51119000, Thermo Fisher Scientific Inc., Waltham, MA, USA).

The protein levels of IFN-γ, IFN-α, interleukin (IL)-1β, IL-2, IL-4, IL-5, IL-6, IL-10, IL-17A and Tumor necrosis factor (TNF) -α in serum were tested using Aimplex kit (Hang zhou CEGER P010041), a multiplex beads-based flow cytometric assay, following the manufacturer’s instructions. The concentrations of IL-4 in the serum samples were further assayed using the ultra-sensitive single molecule array immunoassay on Medivh SX-L platform (ColorTech Biotechnology, Suzhou, China). Samples were incubated with anti-IL-4 capture antibody coated magnetic beads and biotinylated detection antibody to form the immuno-complex. Then beads were loaded into femto-sized array for detection.

### Single-Cell RNA-seq library construction and sequencing

Using a Single Cell 5’ Library and Gel Bead kit (10× Genomics, 1000006) and Chromium Single Cell A Chip kit (10× Genomics, 120236), the cell suspensions (600–1000 living cells per ul as determined by Count Star) were loaded onto a Chromium single cell controller (10× Genomics) to generate single-cell gel beads in the emulsion (GEMs) following the manufacturer’s protocol. Approximately 12,000 cells were added to each channel and approximately 7,000 target cells were recovered. Single-cell RNA libraries were prepared using the Chromium Single Cell 5’ v2Reagent (10× Genomics, PN-1000263) was used to prepare single-cell RNA libraries according to the manufacturer’s instructions. Each sequencing library was generated with a unique sample index. The libraries were sequenced using Illumina platform with a sequencing depth of at least 100,000 reads per cell and 150 bp (PE150) paired-end reads (performed by GENE SHINE, Shanghai).

### Single-Cell Data Processing

The matrices of unique molecular identifier (UMI) were generated for each sample by the Cell Ranger (10× Genomics, Version 4.0.0) Pipeline coupled with human reference version GRCh38 (10× Genomics, Version 3.0.0.) using STAR (version 2.5.1b). Then the expression matrix was analyzed by R software (v.3.6.0) with the Seurat package (v.3.0.0) and DoubletFinder (version 2.0.2) for filtering, data normalization, dimensionality reduction, clustering (Butler *et al*., 2018). The filtered scRNA-seq data were analyzed with the following steps: 1. To remove the cells with low quality, cells with fewer than 500 genes detected and a mitochondrial gene ratio of greater than 10% were excluded. And genes with at least one feature count in more than three cells were used for the following analysis. 2. To remove potential doublets, cells with UMI counts above mean ± 2SD are filtered out. Additionally, we applied DoubletFinder package (version 2.0.2) to identify potential doublets. 3. For each cell, we normalized the gene expression profiles with SCT-transform in Seurat package. 4. Variable genes were selected using the ‘FindVariableFeatures’ function with the default parameters. 5. Variable genes were projected onto a low-dimensional subspace using canonical correlation analysis (CCA) across samples to correct batch effects by using the ‘RunMultiCCA’ functions. 6. A shared nearest neighbor graph was constructed based on the Euclidean distance in the low-dimensional subspace spanned by the selected significant principal components. Cells were clustered using the ‘FindClusters’ function at an appropriate resolution. 7. Cells were visualized using a 2-dimensional t-distributed stochastic Neighbor Embedding (tSNE) algorithm with the ‘RunTSNE’ function. Using the above pipeline, we processed the scRNA-seq data of ∼120k high-quality cells from 24 samples.

### Reference Single-Cell Datasets in comparison

Other than the scRNA-seq dataset generated in this study on the participants of BBIBP-CorV vaccination, we also used the published PBMC scRNA-seq dataset to make meaningful comparison over the immune-responsive signals. The dataset on COVID-19 patients came from GEO accession GSE158055^21^ and the dataset on flu vaccine came from GEO accession GSE148633^22^.

### Cell-type annotation and cluster marker identification

After nonlinear dimensional reduction and projection of all cells into two-dimensional space by tSNE, cells were clustered together according to the similar features. ‘FindClusters’ function was used to define the unsupervised clusters. “FindAllMarkers” function in Seurat was used to find markers for each cluster. Clusters were then preliminarily classified and annotated based on expressions of canonical markers and signature genes of particular cell types. In total, we identified 11 major cell types including CD4^+^ T and CD8^+^ T cells (*CD3D, CD3E, CD3G, CD40LG, CD4, CD8A, CD8B*), γδ T cells (*TRDV2, TRGV9*), B cells (*MS4A1, CD79A, CD79B*), plasma cells (*JCHAIN, MZB1*), NK cells (*NCAM1, GNLY, NKG7*), Monocyte and Dendritic cells (*CD14, FCGR3A, CST3, LYZ, FCER1A, CD1D, LILRA4*) and Megakaryocytes (Mega) (*PPBP, PF4*). We also used SuperCT^33^ and 31 reference cell types [https://github.com/weilin-genomics/rSuperCT_models] to predict the cell types of the total 120k cells. SuperCT predictions were used to for cross-validation. We assigned the final cell types based on the entire cluster rather than single cells. The dominant prediction for the cluster will correct the discrepancy in the same cluster. Additional details of prediction on top of SuperCT 31-type, such as γδT cells, mDC, pDC were assigned by the enriched markers and detailed cell-type boundaries were determined by the Seurat ‘FindClusters’ function with default parameters.

### Identification of differentially expressed genes (DEGs) and function enrichment

The gene expression signals of a certain cell group (by cell type and by sample) were represented by copy number per million UMIs (CPM). See details in the following equation.

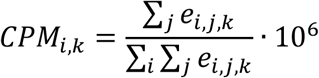

Let *e* be number of unique molecular identifiers (UMIs) of i gene determined by unique molecular barcode in a certain cell j in group k.

The CPMs of each gene were compared using student tests by pairwise or non-pairwise groups. P value < 0.01 was used as a threshold to define the significant differential expression genes (DEGs).

When we compared the differential genes between the post-vaccination time points and the base line samples, we could use pairwise t-test by each individual. When we compare the differential genes across B-R-P sample groups, we could only use independent t-test.

GO and KEGG pathway enrichment analyses on these DEGs identify the biological keywords, that allowed us to summarize the keywords of classical immune responsive signal.

### Transcriptional factor analysis

To identify the potential regulatory transcription factors that dictate the vaccine response, the regulons in major cell types were assessed using SCENIC^27^ with scaled expression matrix as input. The regulons expression profile cluster plots were visualized using the ComplexHeatmap package of R.

### Assessment of the overall signal score of the keywords of immunity

We used cellular modular scores to assess the immune state after infection or vaccination. We selected differentially expressed genes enriched in MSigDB ^34^ to calculate the modular scores. The involved genes for 12 immune-responsive signals were shown in supplemental table3. The modular score of a certain keyword was calculated with ‘AddModuleScore’ function in Seurat with default settings. The keywords signal score of different immune state were analyzed by two-sided wilcox test and all differences with p < 0.05.

### TCR and BCR V(D)J sequencing and analysis

TCR and BCR clonal types were determined using CellRanger V(D)J pipeline(10× Genomics, Version 4.0.0). TCR/BCR diversity was measured as Shannon’s entropy using ‘H’ function in philentropy package:

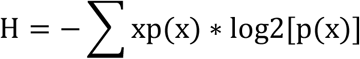

in which the p(x) represents the frequency of a given TCR/BCR clone among all T/B cells with TCR/BCR identified. The TCR/BCR diversity among different times was analyzed by two-sided Student’s t-test and all differences with p < 0.05. We loaded the clonal type information from the CellRanger V(D)J output to the meta data of the seurat objects based on the barcode mapping. The cells belonging to the top 20 TCR and BCR clones were labeled as ‘top-clones’. The gene expression signal and the immune responsive signals were compared between the top-clones and their counterparts using Seurat ‘VlnPlot’ function.

## Data Availability

The data will be online published upon the acceptence of the manuscript by a journal and the agreement by the all authors.

## Acknowledgements

We thank the volunteers from Pu Dong Cohort and Professor Ling Ni at Tsinghua University for their contribution to the study. This work was supported by grants from the National Science Fund for Distinguished Young Scholars (81625002), the National Natural Science Foundation of China (81930007, 81470389, 81500221 and 81800307), the Shanghai Outstanding Academic Leaders Program (18XD1402400), Innovative research team of high-level local universities in Shanghai (SSMU-ZDCX20180200) and Shanghai Municipal Education Commission Gaofeng Clinical Medicine Grant Support (20152209).

## Author contributions

P.J and L.W conceived the need for the article. L.Z, L.Y.Y, H.L, G.X, L.Q, S.T, Z.C.J and Y.A.C recruited volunteers. T.R.Y, C.Y.F, S.J.F, and L.G.Q collected the samples from donors. T.R.Y, H.L.H, C.Y.F, Y.A.C and Z.M.C completed the experiments. L.W, T.R.Y, Z.J.M and L.R.H finished the data analysis. L.W, T.R.Y and C.Y.F drafted the initial version. C.Y.F and T.R.Y provided the production of the flowchart. P.J and L.W put forward many constructive comments for the final version. All the authors read and approved the manuscript.

