## Supplementary material for "Characterizing cellular and molecular variabilities of peripheral immune cells in healthy inactivated SARS-CoV-2 vaccine recipients by single-cell RNA sequencing": Supplementary Figures.pdf

**SUPPLEMENTAL MATERIAL**

**Supplemental Figures**

**Supplemental Figure S1.**

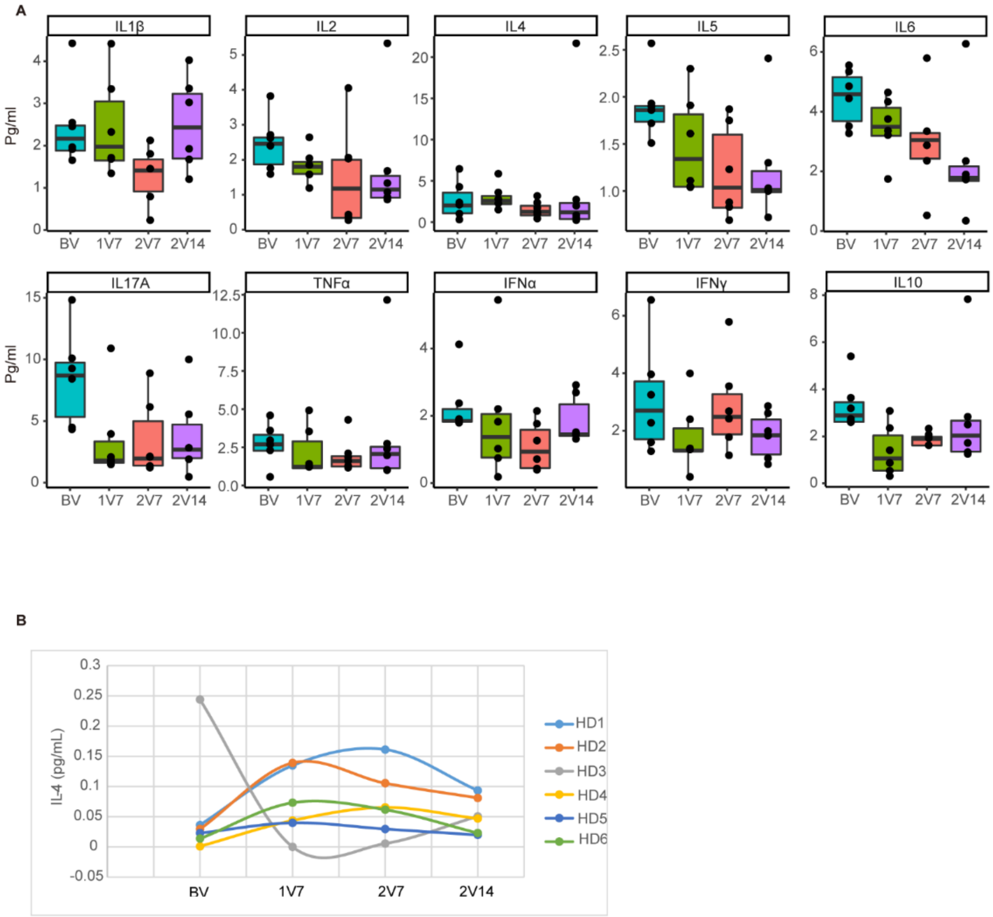


**Figure S1.** The key cytokines detection at the different sampling time.

A. The key cytokines were detected at four time points using BD FortessaX20 platform

B. The IL-4 was examined with single-molecule array technique.

**Supplemental Figure S2.**

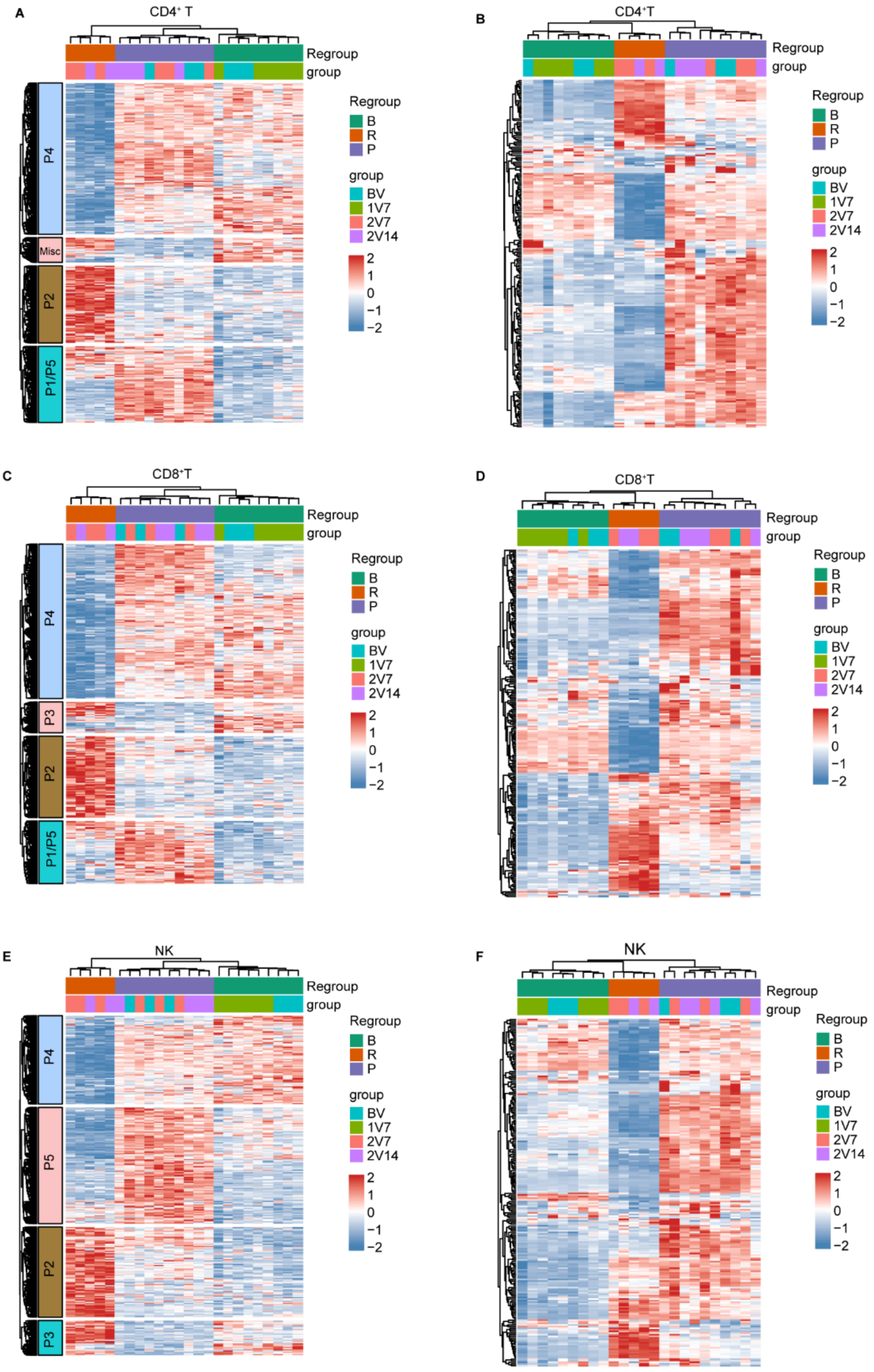

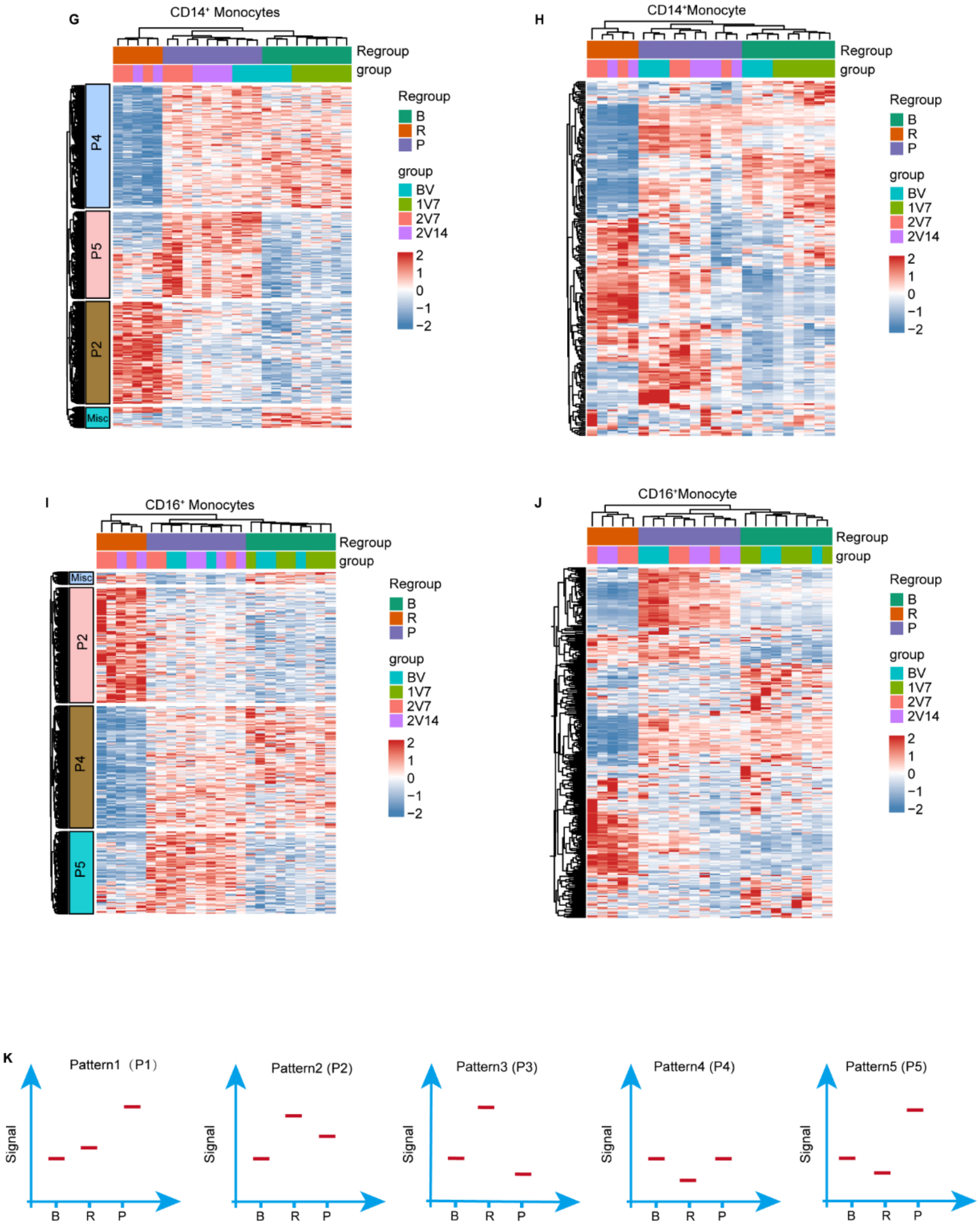
G
CD14⁺ Monocytes
Regroup
group
P4
P5
P2
Misc
Regroup
B
R
P
group
BV
1V7
2V7
2V14
2
1
0
−1
−2
H
CD14⁺Monocyte
Regroup
group
Regroup
B
R
P
group
BV
1V7
2V7
2V14
2
1
0
−1
−2
I
CD16⁺ Monocytes
Regroup
group
Misc
P2
P4
P5
Regroup
B
R
P
group
BV
1V7
2V7
2V14
2
1
0
−1
−2
J
CD16⁺Monocyte
Regroup
group
Regroup
B
R
P
group
BV
1V7
2V7
2V14
2
1
0
−1
−2
K
Pattern1（P1）
Pattern2 (P2)
Pattern3 (P3)
Pattern4 (P4)
Pattern5 (P5)
Signal
B
R
P

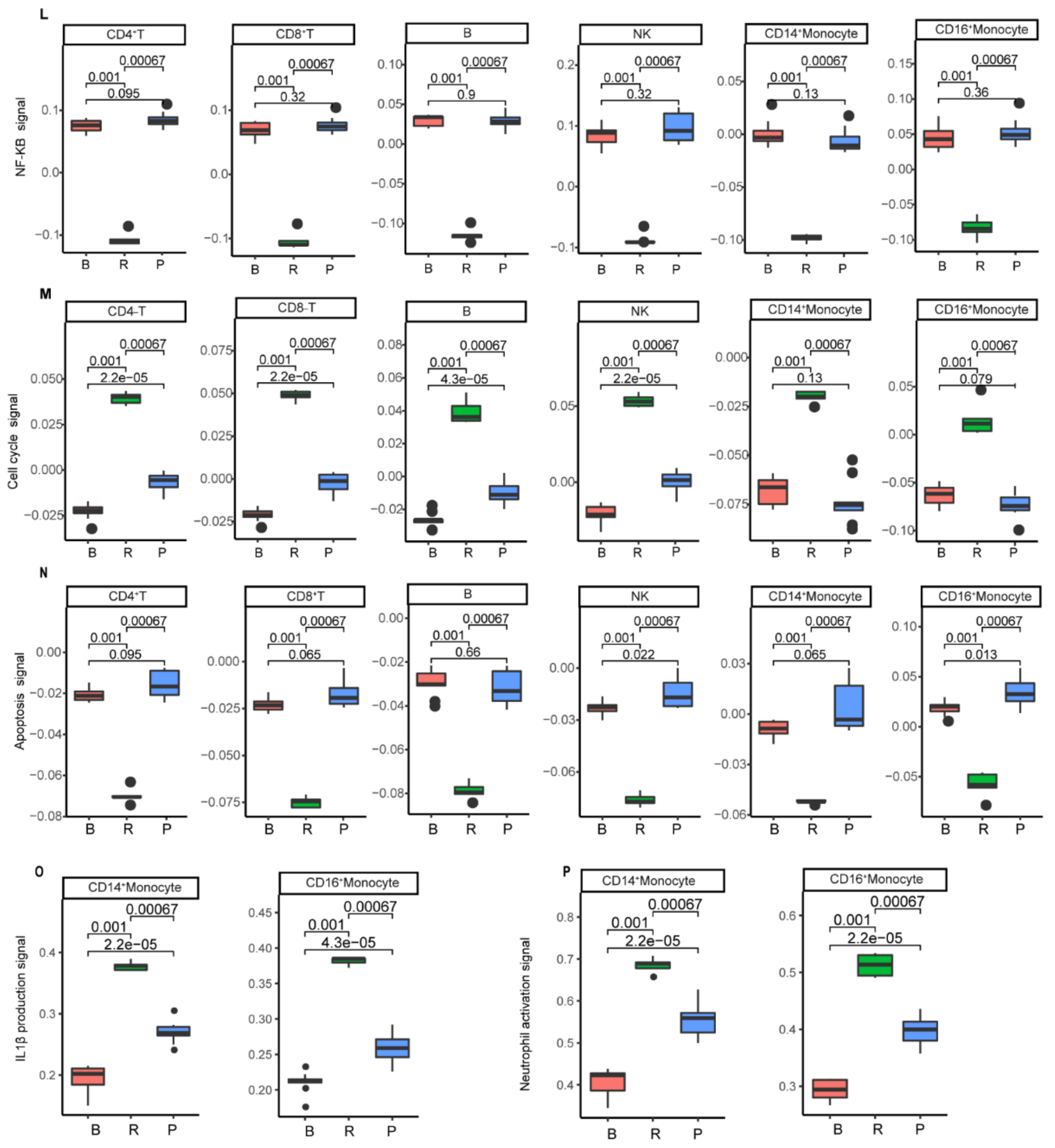


**Supplemental S2**. The DEGs and regulons expression and keywords signals module score of major cell types.

A, C, E, G, I. heatmap of DEGs detected in major immune cell types;

B, D, F, H, J. heatmap of regulons detected in major immune cell types;

K. The simple schematic diagrams of 5 dominant patterns;

L-P. variable immune-responsive signals in the major cell types.

### Supplemental Figure S3.

A

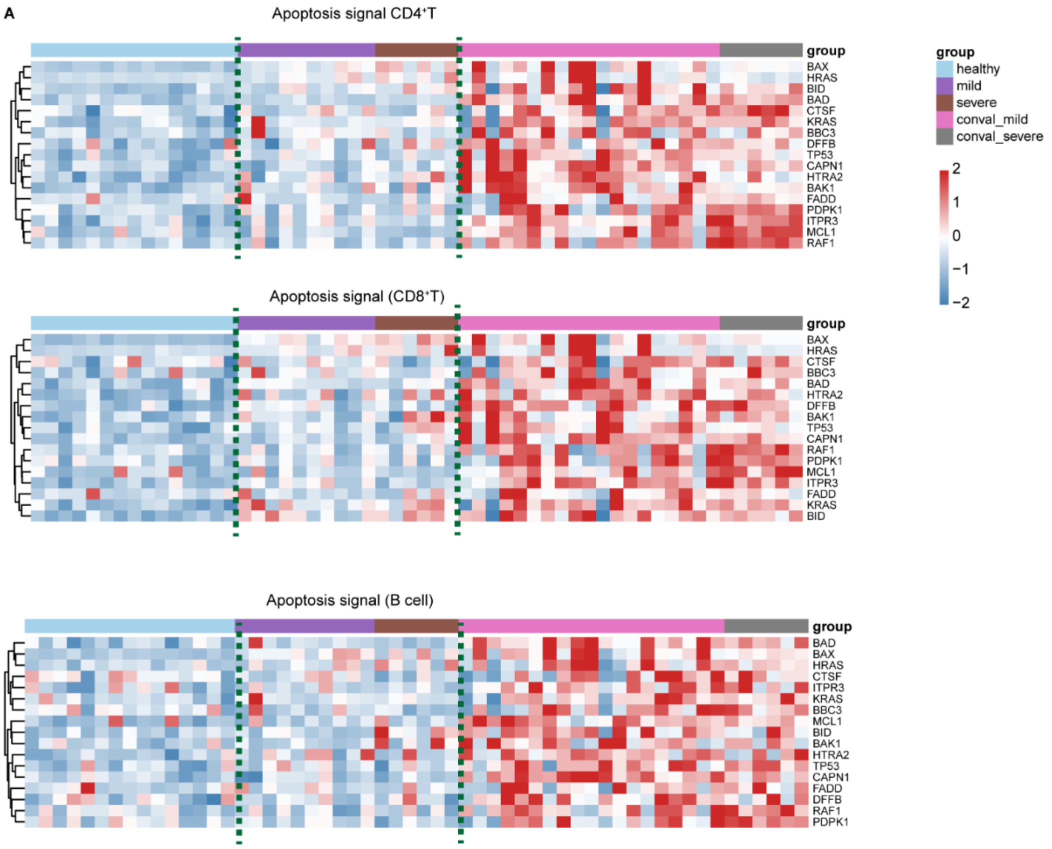


B

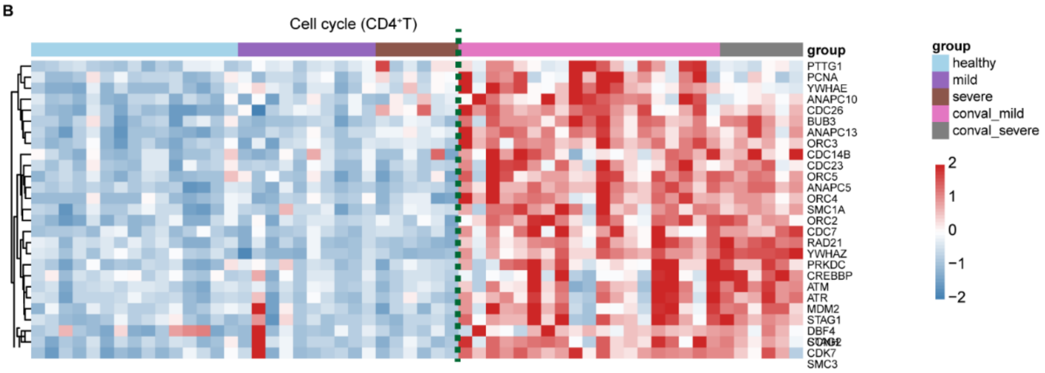

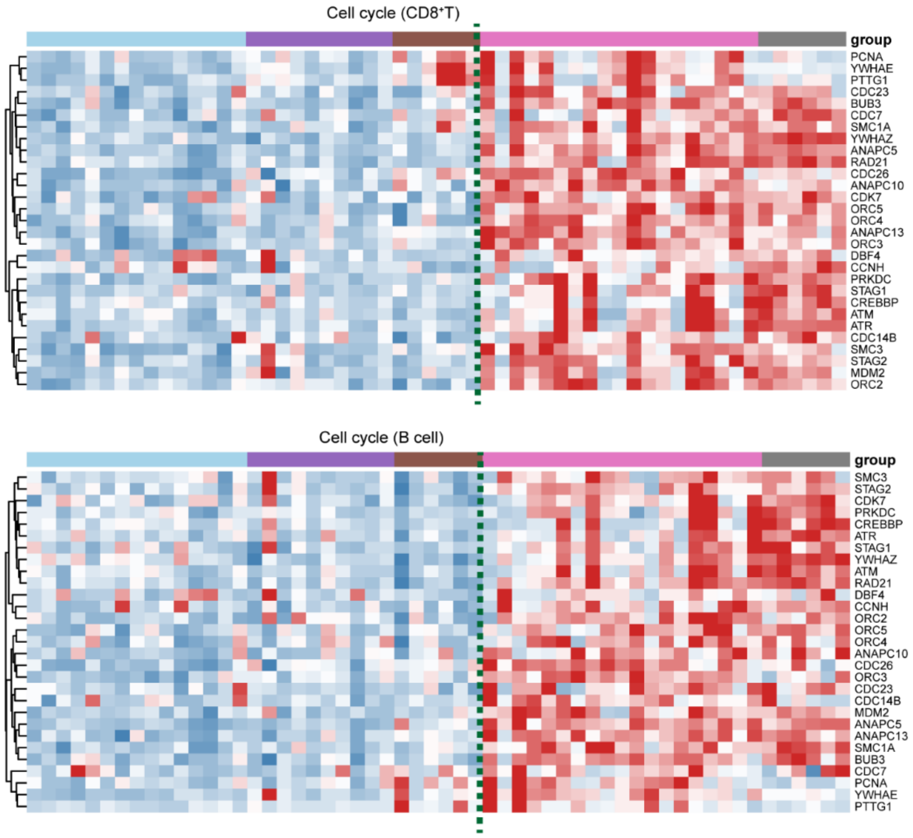
Cell cycle (CD8⁺T)
group
PCNA
YWHAE
PTTG1
CDC23
BUB3
CDC7
SMC1A
YWHAZ
ANAPC5
RAD21
CDC26
ANAPC10
CDK7
ORC5
ORC4
ANAPC13
ORC3
DBF4
CCNH
PRKDC
STAG1
CREBBP
ATM
ATR
CDC14B
SMC3
STAG2
MDM2
ORC2
Cell cycle (B cell)
group
SMC3
STAG2
CDK7
PRKDC
CREBBP
ATR
STAG1
YWHAZ
ATM
RAD21
DBF4
CCNH
ORC2
ORC5
ORC4
ANAPC10
CDC26
ORC3
CDC23
CDC14B
MDM2
ANAPC5
ANAPC13
SMC1A
BUB3
CDC7
PCNA
YWHAE
PTTG1

C

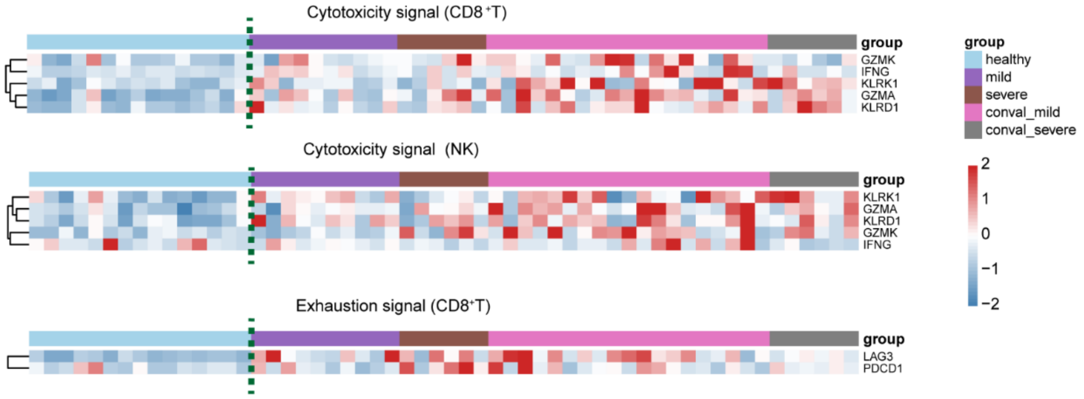
Cytotoxicity signal (CD8⁺T)
group
GZMK
IFNG
KLRK1
GZMA
KLRD1
Cytotoxicity signal (NK)
group
KLRK1
GZMA
KLRD1
GZMK
IFNG
Exhaustion signal (CD8⁺T)
group
LAG3
PDCD1
group
healthy
mild
severe
conval_mild
conval_severe
2
1
0
−1
−2

D

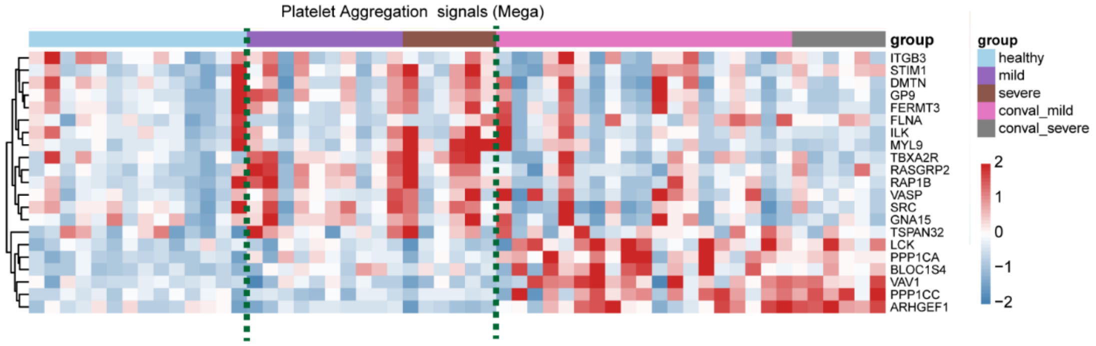
Platelet Aggregation signals (Mega)
group
ITGB3
STIM1
DMTN
GP9
FERMT3
FLNA
ILK
MYL9
TBXA2R
RASGRP2
RAP1B
VASP
SRC
GNA15
TSPAN32
LCK
PPP1CA
BLOC1S4
VAV1
PPP1CC
ARHGEF1
group
healthy
mild
severe
conval_mild
conval_severe
2
1
0
−1
−2

**Figure S3.** Classical immune responsive signal genes of major immune cell types in Covid-19 patients.

A. Apoptosis signal genes of major immune cell types in Covid-19 patients;

B. Cell cycle signal genes of immune cell types in Covid-19 patients.
